# Stakeholders’ views on an institutional dashboard with metrics for responsible research

**DOI:** 10.1101/2021.09.16.21263493

**Authors:** Tamarinde Haven, Martin Holst, Daniel Strech

**Affiliations:** Berlin Institute of Health at Charité – Universitätsmedizin Berlin, QUEST Center, Charitéplatz 1, 10117 Berlin, Germany; Medizinische Hochschule Hannover, Institut für Ethik, Geschichte und Philosophie der Medizin, Carl-Neuberg-Str. 1, 30625 Hannover, Germany

**Keywords:** responsible research, university medical center, dashboard, Open Science, robustness, transparency

## Abstract

**Background:** Concerns about research waste have fueled debate about incentivizing individual researchers and research institutions to conduct responsible research. Instead of looking at impact factors or grants, research institutions should be assessed based on indicators that pertain to responsible research. In this study, we showed stakeholders a proof-of-principle dashboard with quantitative metrics that visualized responsible research performance on a German University Medical Center (UMC) level. Our research question was: What are stakeholders’ views on a dashboard that displays the adoption of responsible research practices on a UMC-level?

**Methods:** We recruited different stakeholders to participate in an online interview. Stakeholders included UMC leadership, support staff, funders, and experts in responsible research. We asked interviewees to reflect on the strengths and weaknesses of this institutional dashboard approach and enquired their perceptions of the metrics it included. The interviews were recorded and transcribed. We applied content analysis to understand what stakeholders considered the Strengths, Weaknesses, Opportunities, and Threats of the dashboard and its metrics.

**Results:** We interviewed 28 international stakeholders (60% German). Overall, interviewees thought the dashboard was helpful in seeing where an institution stands and appreciated the fact that the metrics were based on concrete behaviors. Main weaknesses included the lack of a narrative explaining the choice of the metrics covered. Interviewees considered the dashboard a good opportunity to initiate change and hoped the dashboard could be supplemented with other indicators in the future. They feared that making the dashboard public might risk incorrect interpretation of the metrics and put UMCs in a bad light.

**Discussion:** While the feedback was given specifically to our proof-of-principle dashboard, our findings indicate that discussion with stakeholders is needed to develop an overarching framework governing responsible research on an institutional level, and to involve research-performing organizations.

## Introduction

Concerns about research waste have fueled debate about incentivizing individual researchers and research performing organizations to conduct responsible research (Macleod et al., 2014). One key point was that individual and institutional research performance should be assessed differently (‘DORA’, 2013; Hicks et al, 2015; Ioannidis, 2014). Rather than looking at high impact publications or grants obtained, one should focus on indicators that pertain to responsible conduct of research (Higgison & Munafò, 2016; Moher et al., 2018; Moher et al., 2020). This holds true for individual researchers of all career stages (Flier, 2017), as well as for research performing organizations, that can themselves be subject to dysfunctional incentives (Anderson, 2019; Bagioli et al., 2019).

Research performing organizations, in our case, University Medical Centers (UMCs), play a key role in fostering responsible research (Begley et al., 2015; Strech et al., 2020; Bouter, 2020). They can put out relevant policies (e.g., for data sharing), provide critical infrastructure (e.g., for coordinating clinical trials), and reward responsible research more generally (Rice et al., 2020). But this requires awareness about responsible research on an institutional level, and commitment from UMCs to make this a priority (Bouter, 2018; Forsberg et al., 2018).

In this study, we showed stakeholders a proof-of-principle dashboard with quantitative metrics that visualized responsible research performance on a UMC level (e.g., for a given UMC, what percentage of its trials is prospectively registered?). The metrics included pertained to responsible research practices such as registration and reporting of clinical trials (inspired by work from Goldacre et al., 2018; Wieschowski et al., 2019), robustness in animal research (e.g., randomization and blinding, see Macleod & Mohan, 2019), and Open Science (including Open Access, Data, and Code, see Serghiou et al., 2021). Our research question was: What are stakeholders’ views on a dashboard that displays the adoption of responsible research practices on a UMC-level?

## Methods

### Ethical approval

The ethical review board of the Charité – Universitätsmedizin Berlin reviewed and approved our research protocol and materials (#EA1/061/21).

### Participants

When discussing the adoption of responsible research practices on an institutional (UMC) level, we distinguish four broad stakeholder groups: 1) UMC leadership (e.g., deans, heads of strategy, heads of pan-UMC organizations), 2) support staff (e.g., policy makers, librarians, heads of core facilities (including 3R centers and clinical research units)), 3) research funders, and 4) experts in responsible research assessment. Participants were recruited through our own networks, snowballing, or through internet searches (cold calling). We used purposive sampling, combined with snowballing (i.e., following up on potential stakeholders suggested by interviewees) and invited a total of 49 potential participants.

### Procedure

Participants first received an invitational e-mail with a link to the information letter, informed consent form, and protocol (see appendix 1). We sent one reminder a week after. When participants agreed to participate, we scheduled an appointment for an online interview through Microsoft Teams. Participants received a link to the dashboard and a tutorial explaining the dashboard (see <u>here</u> and appendix 2). They signed the informed consent form prior to the interview.

We conducted the interviews online between April and June 2021. Interviews lasted between 30 and 50 minutes. One team member led the interview (TH (Dr), MH (PhD candidate) or DS (Prof), who are all trained in qualitative research methods), whilst another team member observed and made notes on the interview’s process and its content. After a brief introduction of the interviewer and the interviewee, interviews were conducted using a topic guide (semi-structured) that was based on a literature review and various internal discussions (see appendix 3). The topic guide was pilot tested (*n* = 3) for comprehensibility using cognitive interviewing (Beatty & Willis, 2007). After each interview, the team got together for reflection (peer-debriefing). If they wanted, interviewees received a brief written summary of the interview (member check) with the option to comment or correct. The interviews were transcribed by a transcription company under a data processing agreement. We used the COREQ checklist to write up our findings (Tong et al., 2007). For a more elaborate description of our procedure, see study protocol (https://osf.io/z7bsg/).

### Data analysis

We applied content analysis that involved deductive and inductive elements (Elo & Kyngäs, 2008). We structured stakeholders’ views by looking for Strengths, Weaknesses, Opportunities, and Threats (SWOTs), see Table 1. Subcodes within each part of the SWOT were derived from the data. These subcodes pertain to the general approach of an institutional dashboard and responsible metrics, i.e., suggestions for improvement by interviewees that were specific to the proof-of-principle version are not featured in the results where they have been implemented successfully (e.g., the dashboard interviewees saw displayed a sample of publications, the final dashboard that will be published shortly includes all publications).

**Table 1.** SWOT definitions

|  |  |
| --- | --- |
| **Strengths** | Characteristics inherent to the current dashboard approach or the metrics included that are considered valuable for visualizing institutional performance in terms of responsible research |
| **Weaknesses** | Characteristics inherent to the current approach that could be considered disadvantages, areas that need improvement for the dashboard and that are within the realm of the internal environment |
| **Opportunities** | Potential use cases of the dashboard that could increase its chances of successful uptake, as well as additions to the approach or the metrics that – when implemented – would improve the chances of stakeholder wide-uptake of visualizing institutional performance in terms of responsible research |
| **Threats** | Characteristics in the external environment that could undermine implementation of the dashboard approach or others measures to visualize institutional performance in terms of responsible research |

Our analysis consisted of three phases. First, two team members (TH and MH) read and coded 5 interviews independently using MAXQDA 2020 (Release 20.3.0, VERBI GmbH, Germany). They then each proposed a code tree, exchanged these, and resolved discrepancies through discussion. With this revised code tree, the two team members coded another 5 interviews. This process was repeated until saturation was established (Fusch & Ness, 2015). The code tree that resulted was then presented to the full research team (together with the interviews). Reviewing the final interviews resulted in minor modifications to the wording of the code tree only (appendix 4), which formed the basis of our results.

## Results

### Demographics

We invited a total of 49 stakeholders, 28 of them agreed to an interview (response rate: 57%, one interview was conducted with two participants). From these 28 stakeholders, 89% came from our own networks or the networks of our collaborators, and 11% were suggested through snowballing. The demographics of our interviewees appear in Table 2.

**Table 2.** Demographics of interview participants

| **Stakeholder group** | **#** |
| --- | --- |
| *UMC leadership* | 4 |
| *Support staff (including librarians, policy makers, CRU staff, 3Rs staff)* | 14 |
| *Funders* | 5 |
| *Responsible research experts* | 5 |
| **Gender** |  |
| *Male* | 22 |
| *Female* | 6 |
| **Nationality** |  |
| *Germany\** | 17 |
| *Europe (besides Germany)* | 10 |
| *North America* | 1 |
| **Total** | 28 |
| *\* One interview was conducted in German.* |  |

Below we describe the most important SWOTs according to our interviewees. We identified 3 strengths, 3 main weaknesses, 6 opportunities, and 4 main threats, see Figure 1. Each subtheme is illustrated with quotes, see Table 3.

**Figure 1.**
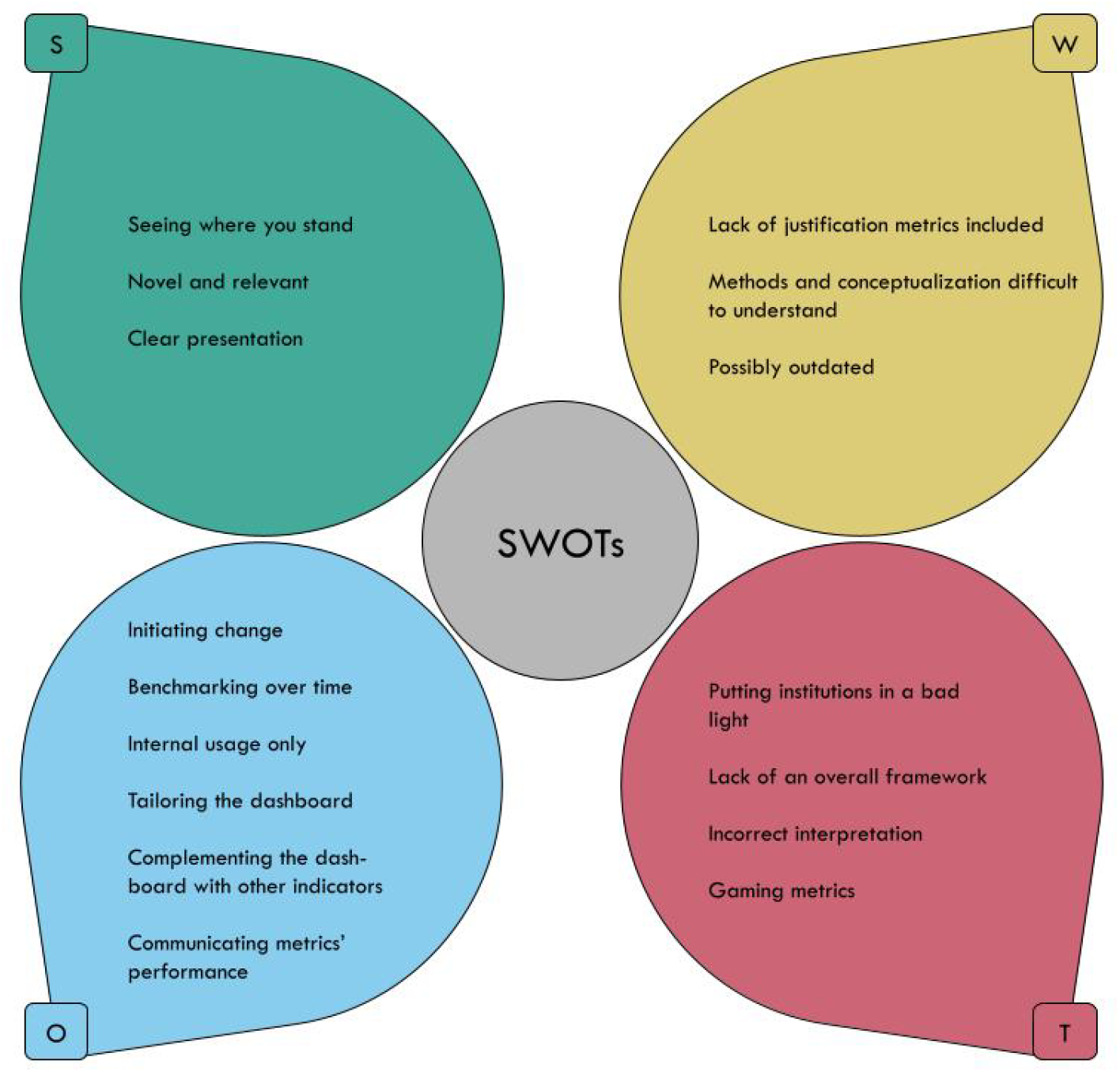
Overview of SWOTs regarding institutional dashboards with metrics for responsible research

**Table 3.**
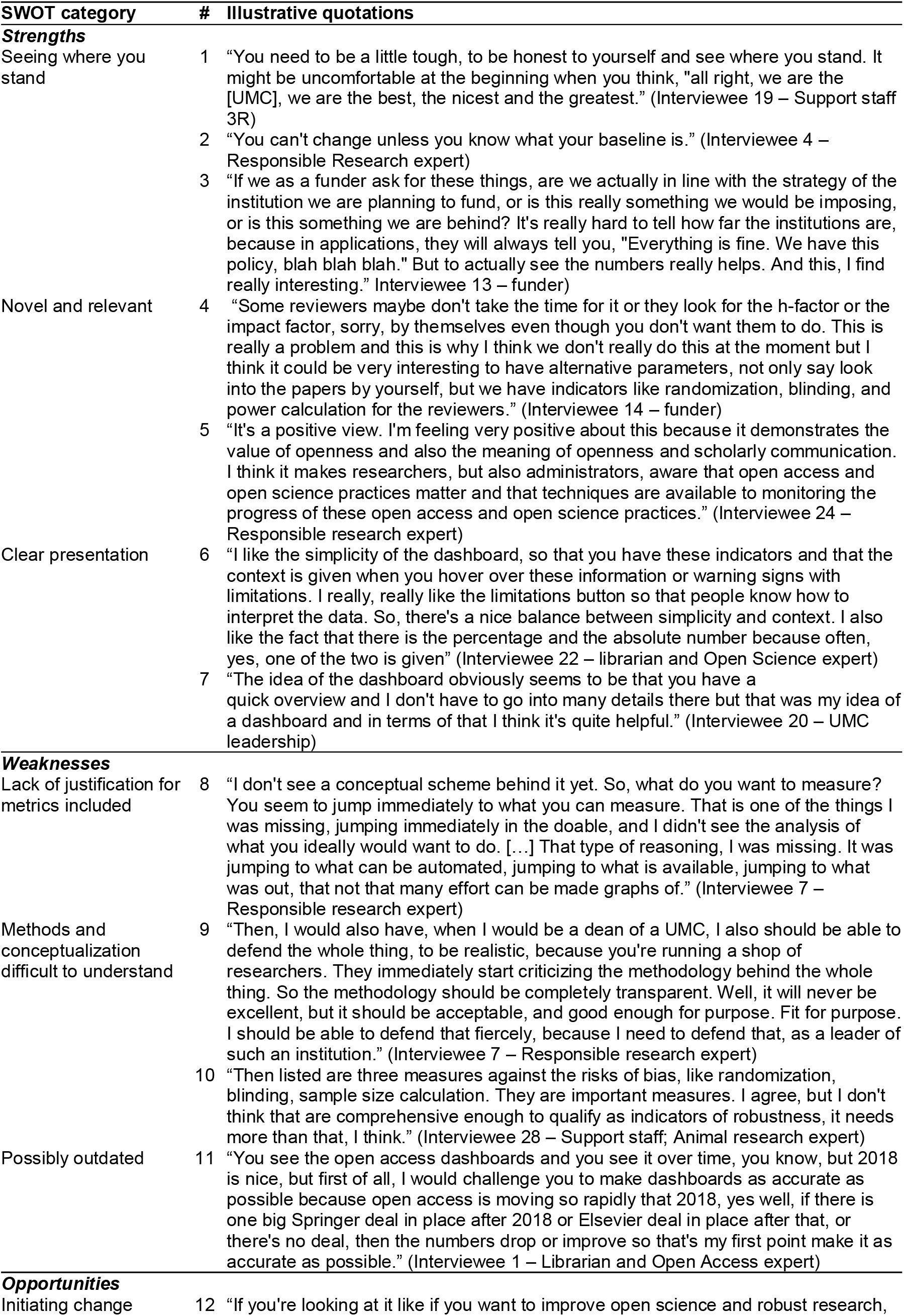
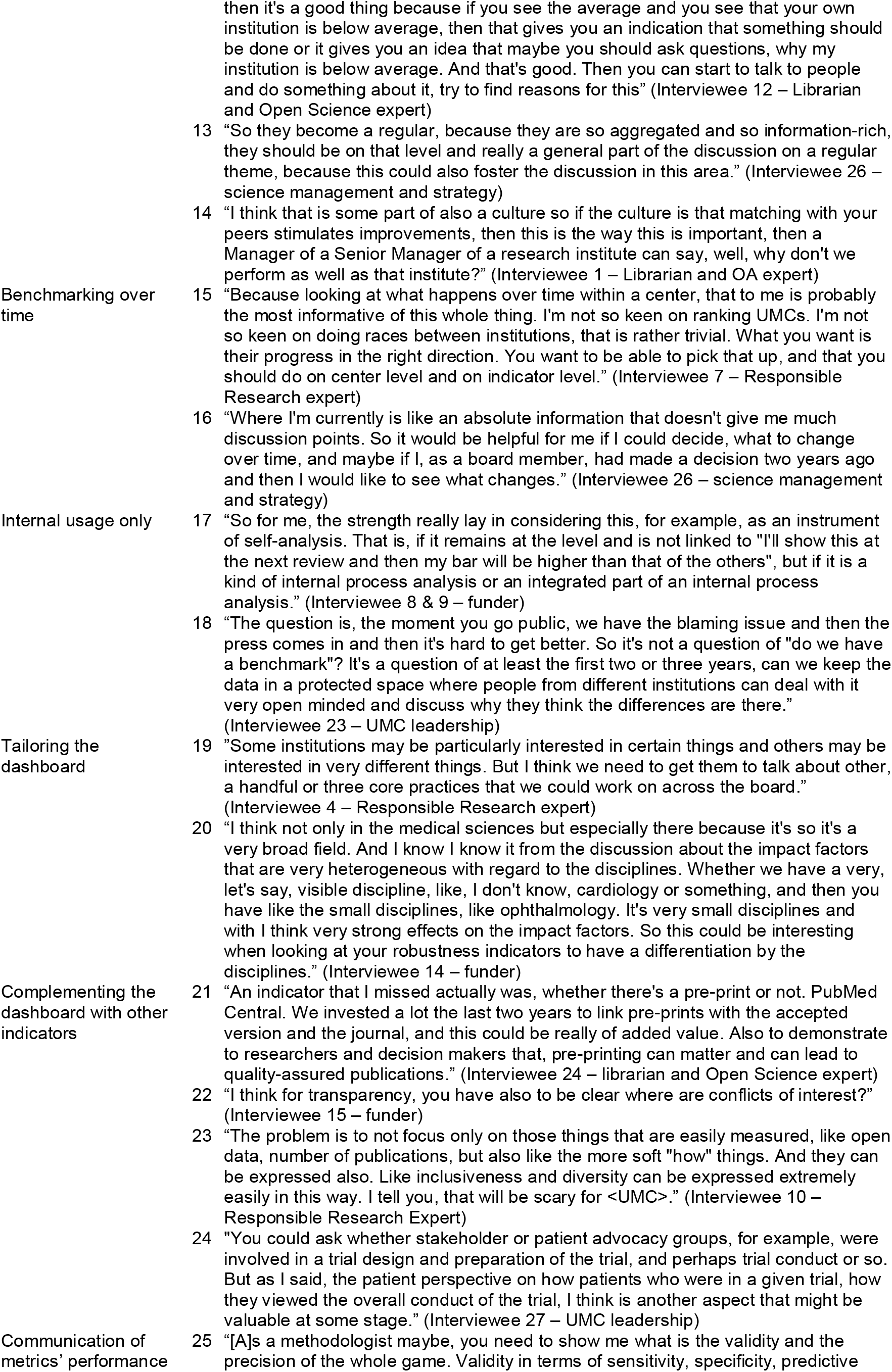
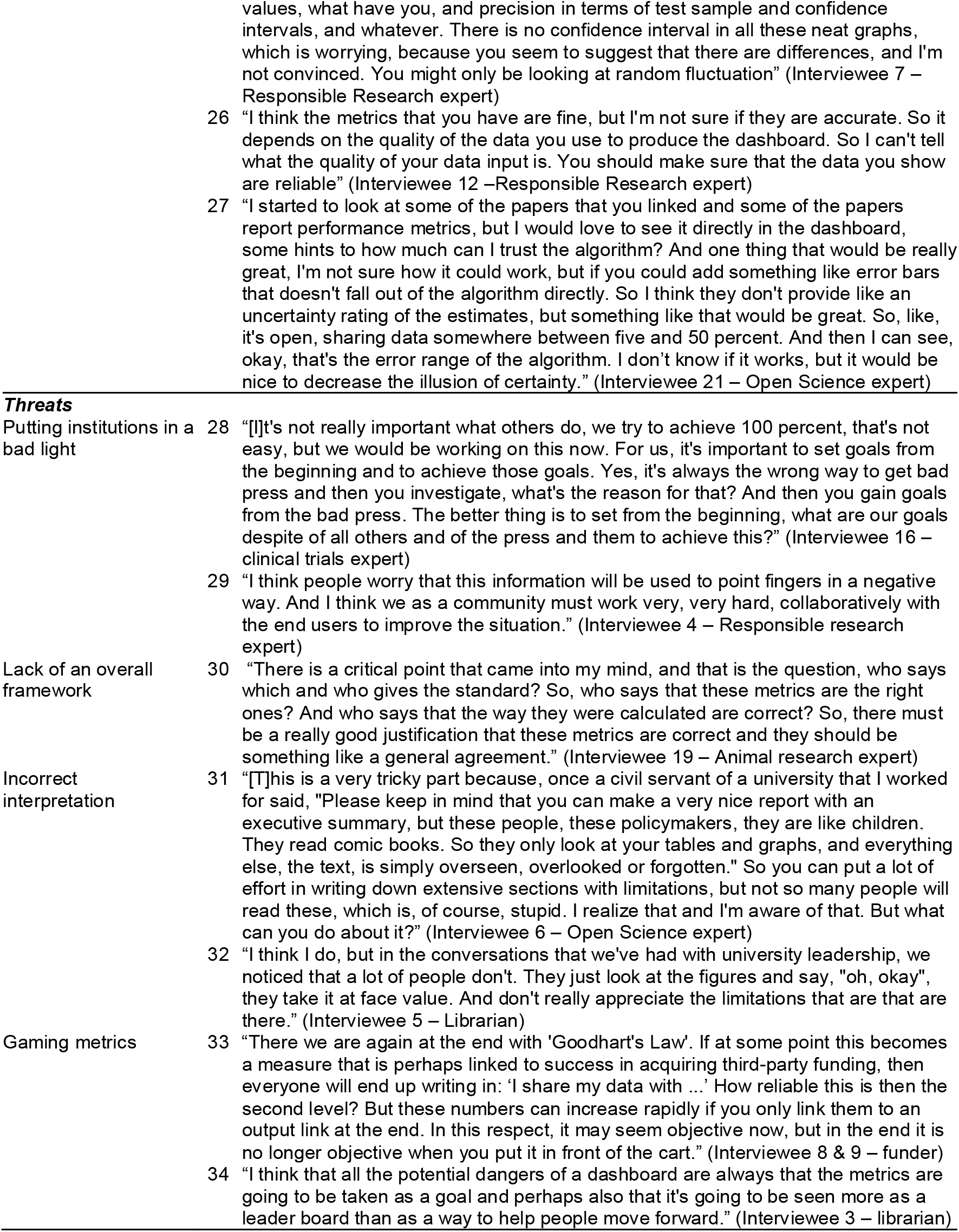
Illustrative quotes per theme.

### Strengths

#### Seeing where you stand

Overall, interviewees were pleased to see that the dashboard focused on specific measures that relate to concrete behaviors. Many interviewees thought the dashboard was helpful in showing institutions where they stand when it comes to responsible research practices. The dashboard approach allowed for creating a baseline, which was considered essential before talking about change or improvement on the metrics. Some expressed curiosity about the extent to which the dashboard would match internal data on self-evaluation processes.

#### Novel and relevant

Various interviewees indicated this was the first time they saw such a dashboard. They thought it fulfilled an important need and some interviewees felt these measures could serve as an alternative to publication output, journal impact factors, third-party funding, or other widespread metrics of institutional performance. Many interviewees considered the included topics timely, as they related to ongoing discussions in the field regarding responsible research and contemporary science policy.

#### Clear presentation

Many interviewees indicated they appreciated the transparent overview. They liked the interactive features of the dashboard (e.g., switching between percentages and absolute numbers, hover-over fields with limitations) and appreciated the fact that it gave them a good overview whilst not overburdening the viewer with information.

### Weaknesses

#### Lack of justification for metrics included

Several participants indicated that they missed an overall narrative that would justify the choice of metrics included. In addition, it was unclear to some interviewees how the metrics related to each other, or to the quality of research. This made it hard for participants to understand the purpose of the dashboard. By presenting these metrics, it seemed as if these metrics were the standard metrics for institutional assessment, and various participants indicated that they did not consider that accurate.

#### Methods and conceptualization difficult to understand

Several interviewees expressed difficulty in interpreting the methods by which the metrics were compiled and the limitations inherent to these methods. This included the fact that the metrics may change over time, e.g., the numbers for Green Open Access might change once more publications are made available. Various participants stressed that the way particular metrics were operationalized was unclear. Finally, participants found it difficult to understand the metrics’ denominators, indicating it was unclear whether all metrics pertained to the same dataset or to different (parts of the) dataset(s), and whether the dashboard used all relevant available data sources (e.g., all eligible study registries), to provide a less biased measurement.

#### Possibly outdated

Some participants were concerned that metrics on robustness of animal research or Open Science stemming from publications in 2018 would present an outdated picture. They pointed to various developments in those areas in recent years and interviewees expected that publications from 2019 and 2021 would show higher robustness and Open Science scores. Although the fact that the data were taken from 2018 was a feature of the proof-of-principle dashboard, interviewees’ concerns underscore the need to continuously integrate new information into the dashboard, ideally in an automated manner.

### Opportunities

#### Initiating change

The dashboard was considered a good tool to start an evidence-based conversation on how to improve responsible research practices and about a more holistic evaluation of research institutions, i.e., assessing different institutional facets and not merely citations or impact factors. Some interviewees remarked that the dashboard could provide more information about how one could induce change, and aid in the development of concrete interventions. The dashboard would then, in their view, need to be presented to organizational leadership on a regular basis to fuel a discussion about efforts to improve. In addition, participants thought the dashboard could promote collective improvement efforts (or ‘healthy competition’) that would enable institutions to learn from each other.

#### Benchmarking over time

Several interviewees stressed that the dashboard should include the possibility to benchmark oneself over time. Whereas this was possible for some indicators, many interviewees believed that having this data over time for all metrics could help institutions to evaluate whether interventions to improve their performance on particular metrics were successful.

#### Internal usage only

When thinking about how to prevent possible detrimental consequences of benchmarking, many interviewees stressed that it would be most responsible if the dashboard would be used for internal purposes only. The dashboard’s information would not be accessible to anyone outside the institution, except if the institutional leadership chose to publicize some information. Some interviewees suggested that the dashboard should go public with a 2- or 3-year delay, which would give institutions the opportunity to improve first.

#### Tailoring the dashboard

Another opportunity that various interviewees raised was tailoring the dashboard to the institutional strategy. This would mean that different institutions would have different dashboards with different metrics, allowing institutions to create focus areas for a specific timeframe (e.g., in 3 years’ time, at least 60% of our publications should have open data). Related to this, some interviewees remarked that it would be helpful to link to more fine-grained levels of information, e.g., on a particular center within the research institution or a research field level. They remarked that they would appreciate the opportunity to correct or at least comment on the numbers (e.g., providing a reason why an institution performs badly on trial reporting), or to withdraw from the dashboard.

#### Complementing the dashboard with other indicators

Various interviewees discussed additional metrics they thought might be included the dashboard. Two of the most often mentioned suggestions were preprints and conflict of interest statements. Interviewees involved in clinical trials thought a metric on the overall number of clinical trials conducted could provide useful context. They also suggested metrics related to patient engagement in clinical research. In relation to animal research, some interviewees thought a metric for preregistration of animal studies could be useful, while others mentioned the choice of the model system as an important metric for the generalizability and translation of research. More broadly speaking, various interviewees mentioned metrics related to diversity, such as the percentage of female researchers or female principal investigators at a particular institution, or metrics related to the societal value of research, such as uptake of research by the media, policy documents, clinical guidelines, or patents.

#### Communicating metrics’ performance

Different interviewees remarked that they missed statistics on metrics’ performance. They would like to see uncertainty indicators such as confidence intervals, so that they could assess when an institute would perform better or worse than all institutions together. They also asked for numbers on the sensitivity and specificity of the sampling and classification approaches.

### Threats

#### Putting institutions in a bad light

Many interviewees feared that making the dashboard public in its current form would put institutions in a bad light. Whereas they thought some form of benchmarking could be helpful, they worried that the dashboard could be used as a tool to name and shame institutions, which could in turn have detrimental consequences for funding or for an institution’s reputation among the public. Some interviewees also mentioned that institutions should not be blamed for not meeting a metric’s requirement in the absence of sufficient infrastructure in place (e.g., a data sharing platform) that would allow institutions to comply. A few interviewees also pointed out that there is no inherent need for a comparison, since it is already self-evident that institutions should do their best to score high on these metrics.

#### Lack of an overall framework

Some participants pointed out that the metrics seemed normative but were not backed by an overall legal or regulatory framework. This made participants question who decides what good metrics for responsible research are. Without such a consented framework, participants feared that the dashboard would not be accepted as a viable complementary or alternative approach to evaluate research institutions.

#### Incorrect interpretation

Various interviewees were concerned that that the metrics would be misinterpreted, whether intentionally or unintentionally. Especially under precious time, stakeholders might not appreciate the metrics’ limitations and instead take them at face value. This might result in inflating minor differences and risk the resource-heavy implementation of policies based on little understanding of the metrics. Additionally, some interviewees wondered whether other German UMCs were the right comparator, as there are large differences between the institutions.

#### Gaming metrics

Finally, various interviewees thought that the dashboard and the indicators therein could be gamed. Researchers at the respective institutions might feel that they must score high on these metrics and might get creative in the ways to score high, without living up to the ideals included in the respective metrics. More generally, some interviewees felt that boiling down these important issues to numeric indicators was not without risks.

## Discussion

We enquired stakeholders’ views on a dashboard that displayed the adoption of responsible research practices on a UMC-level. Overall, interviewees were positive about the dashboard approach, indicating that it allowed them to “talk reality”. Various interviewees missed a justification for why these specific metrics and no other potentially relevant metrics for responsible research were included. Some interviewees expressed difficulty in understanding how the metrics were derived from the data. Different interviewees believed the dashboard could be instrumental in sparking behavioral change on an institutional level and hoped the dashboard would include more diverse metrics in the future. The main fear among interviewees was that making the dashboard public would risk metrics being taken at face value and could put UMCs in a bad light.

### Contextualization

#### Overarching frameworks

Various interviewees talked about missing an overall framework to fit this dashboard. There is a variety of frameworks available that are all applicable to the topic of responsible research practices. Broad examples include the TOP guidelines (Nosek et al., 2015) that include, beyond data and code transparency, study preregistration and openness about materials, and focus on what journals incentivize. Another broad example, this time focused on individual researchers, are the Hong Kong Principles (Moher et al., 2020), where explicit attention is paid to peer review and, more recently, diversity (Moher et al., 2021). More specific frameworks include ARRIVE for animal research (Kilkenny et al., 2010), or the Cochrane Risk of Bias tool for clinical trials (Higgins et al., 2011).

Some of our metrics find their origin in regulatory frameworks, such as the mandated registration of drug trials in EudraCT (European Commission, 2012). In addition, the World Health Organization has specified that results of clinical trials should be published in a relevant journal within two years of trial completion (WHO, 2015; 2017). The Declaration of Helsinki (World Medical Association, 2013) calls for all research involving human subjects to be prospectively registered and published.

That said, no framework for responsible research seems broad enough or has received universal or consented support. It is questionable whether such a framework will be developed, and it raises the question of who should oversee its development. A potential candidate would be the UNESCO recommendations of Open Science (UNESCO, 2021). An alternative approach could be involving a group of UMCs in a consensus-building procedure to find a set of indicators for responsible research that UMCs agree upon (see Cobey et al., 2021).

#### Not making public data public

Several interviewees were skeptical of making the dashboard public and allowing everyone to see how UMCs were doing in terms of responsible research compared to the average, or even compared to each other. This is interesting, as the data that the metrics are based on comes from public databases (e.g., clinicaltrials.gov or PubMed), and most of the tools used to create the metrics are publicly available as well. What then is the ideal level of openness or publicness of this information? We are presented with the dilemma that disclosing the dashboard to a select group of institutional leadership would be safer but would risk that nothing happens. Making the dashboard public would be more likely to spark discussion but could harm the reputation of some institutes. The sensitivity of the topic underscores the need to collaborate closely with the respective research performing organizations when implementing a dashboard for responsible research.

#### Dashboards for institutions

We witnessed a surge in dashboards for responsible research and related topics in recent years. Some have a broad focus, such as the European Commission Open Science Monitor or the French Open Science Monitor that display broad disciplinary fields (Jeangirard, 2019; European Union, 2021). Others focused on (and ranked) individual researchers in terms of transparency, such as the Curate Science Transparency leaderboard (Curate Science, 2021). Some of our interviewees questioned whether an entire UMC is the right level for developing such a dashboard. It is interesting in this context that the DFG’s revision of the Guidelines for Safeguarding Good Research Practice (DFG, 2019) includes explicit chapters on responsibilities for heads of research organizations. We believe that a dashboard approach could support heads of research organizations to empirically monitor the implementation of the revised code of conduct.

### Strengths and weaknesses of the study

This is the first comprehensive map of stakeholders’ views on a dashboard that visualizes UMC performance using metrics for responsible research. We interviewed both stakeholders within the German biomedical research landscape that are familiar with existing UMC incentives, as well as those that worked at universities outside Germany who could provide an international perspective.

There are some limitations to acknowledge. First, the stimulus we used regarded a proof-of-principle dashboard and therefore some SWOTs may be specific to our dashboard and less generalizable. We tried to mitigate this by using interviewees’ feedback on our dashboard to highlight broader issues, such as the lack of an overall consented framework governing responsible research on an institutional level.

Second, stakeholders who are generally skeptical of the idea to evaluate UMCs differently might decline the invitation to participate in a study like ours. Although some stakeholders were very critical, the views presented in this paper might be rosier than the average stakeholder. We tried to invite stakeholders from outside our network, especially German UMC leadership (*n* = 15), but despite various reminders, they were not willing to participate or did not reply.

## Conclusion

We described what stakeholders considered the strengths, weaknesses, opportunities, and threats of a dashboard displaying the adoption of responsible research practices on an institutional level. Stakeholders appreciated the focus on behaviors that allowed them to see where a UMC stands but pointed to the lack of a justification for the metrics included. They feared institutions might be put in a bad light, underscoring the need for close collaboration with research institutions when implementing alternative approaches to evaluate research institutions.

## Data Availability

Interview transcripts are not publicly available as it would infringe upon participants' privacy.

## Acknowledgements

We would like to acknowledge Delwen Franzen, Nico Riedel, Benjamin Gregory Carlisle and Maia Salholz-Hillel for their work in designing the dashboard and feedback on the interview topic guide. We are grateful for the interviewee suggestions from Miriam Kip, Evgeny Bobrov, Simon Krupka, Sarah Weschke, Ulrich Dirnagl and Stephanie Müller-Ohlraun. We would also like to acknowledge Lena Woydack, Constance Holman and Tim Neumann for participating in the pilot interviews and helping us to refine our interview questions. Finally, we are thankful for all interviewees that gave us their time and their views.

## Appendices

1. Communication to interview participants (i.e., invitational e-mail, information letter, informed consent, further correspondence)
2. Proof-of-principle dashboard in pictures and tutorial: https://www.youtube.com/watch?v=VDdljq5zI9E
3. Topic guide
4. Code tree

## Notes

**Conflicts of Interests:** The authors build part of the QUEST Center that developed the dashboard and endorse the increased application of measures for robust and transparent science.

### Competing Interest Statement

The authors build part of the QUEST Center that developed the proof-of-principle dashboard and endorse the increased application of measures for robust and transparent science.

### Funding Statement

This work was funded by the German Federal Ministry of Education and Research (BMBF 01PW18012). The funder had no role in the study design, data collection and analysis, decision to publish, or preparation of the manuscript.

### Author Declarations

The ethical review board of the Charite - Universitaetsmedizin Berlin reviewed and approved our research protocol and materials (#EA1/061/21).

