## Appendices for "Stakeholders’ views on an institutional dashboard with metrics for responsible research"

**Appendix 1.** Communication to interview participants (i.e., invitational e-mail, information letter, informed consent, further correspondence)

Dear …,

We would like to invite you to participate in an interview to gain your expert view on the strengths and weaknesses of a new type of **dashboard** that displays the adoption of **measures for robust and open science** at a specific University Medical Center (UMC).

We developed such a proof-of-principle dashboard in the BMBF-funded project BRAVO. The dashboard displays UMC-specific information on a) Open Science, b) the registration and reporting of clinical trials, and c) the reporting of robustness measures in animal research. We are now seeking feedback on this dashboard from various stakeholders such as funders, scientists, library staff, and UMC leadership.

TAILOIRED TO FUNDERS:
Given your role as a funder in incentivising responsible research practices, we would very much appreciate your feedback on this dashboard.

TAILOIRED TO RCR EXPERTS:
As your work on [paper] has really shaped the debate on research assessment, we would very much appreciate your feedback on this dashboard.

TAILOIRED TO SUPPORT STAFF:
In your role at the library and expertise in assessing how your institute is performing research-wise, we would very much appreciate your feedback on this dashboard.

TAILOIRED TO UMC LEADERSHIP:
In your role as [dean/department lead/etc.], we would very much appreciate your feedback on this dashboard, as it was created to help German university medical centres visualise the adoption of responsible research practices at their institution.

By participating in this interview study, your views may help inform future policy making on research assessment. The interview will focus on the following core questions:

- What would you consider are the strengths and weaknesses of this dashboard approach?
- Which metric(s) would you find most informative to support decision making, and why?

Could you please let us know by replying to this email if you would be willing and available to participate in an (online) interview? We will then schedule an interview time that fits your calendar.

If you are willing to participate, you will be sent a link to the proof-of-principle dashboard as well as a short explanatory tutorial. The interviews will be conducted online between March and May and last between 30-45'. We offer an allowance of €150 for your time and effort. Please see the attached information letter for more background information on the study, a link to the study protocol, as well as detailed information on confidentiality and data protection.

If you know any other suitable colleagues who would be interested, please share their name with us. Should you have any questions, please don't hesitate to contact me, Tamarinde Haven, via.

Thank you very much for your consideration, on behalf of Prof. Daniel Strech and the BRAVO study team at the BIH QUEST Center,

Tamarinde Haven

**DASHBOARD VISUALISING METRICS FOR ROBUST AND OPEN RESEARCH**
***Study information and consent form***

Current indicators for scientific performance are citation metrics and secured third party funding. In order to increase value and reduce waste in the biomedical sciences^[1]^, it has been argued that research institutions should move away from traditional metrics of research evaluation and towards metrics that reflect responsible research practices^[2]^, such as the timely reporting of clinical trial results and open access publications^[3]^.

The BMBF funded project ‘BRAVO’ developed a dashboard (in a proof-of-principle version) that allows UMCs to visualize the adoption of responsible research practices at their institution by means of ‘metrics’ that relate to open science, the registration and reporting of clinical trials, and the robustness of animal studies. This proof-of-principle version serves to illustrate the main features of the dashboard, which is still in development.

In order to understand how stakeholders view the Strengths, Weaknesses, Opportunities, and Threats (SWOTs) of this dashboard approach, we want to conduct in-depth interviews with these main aims:

1.	To determine and better understand stakeholders’ views on this proof-of-principle dashboard displaying metrics (that visualize)/as indicators of responsible research practices

2.	To better understand participants’ views on these different metrics intended to display the adoption of responsible research practices on a research-institute level

**How your input will be used**

The interviews will be recorded with a recording device or software and will be transcribed by members of the study team of external employees of a transcription service. External personnel will not receive any information about participants’ identity or institution; the audio file will be destroyed after transcription, in accordance with data protection laws. The results will be used in a scientific paper, where we will illustrate our findings with quotes from the interview. These quotes are anonymized, but you might be able to recognize your own citations. The goal of the study would be an overview of the strengths, weaknesses, opportunities and threats associated with this dashboard approach and its associated metrics. You can find our full study protocol here: <https://osf.io/ny8az/>.

Prof. Dr. Dr. Daniel Strech

Deputy Director QUEST Center

16. Sep. 2021

**Voluntariness**

Participation in the interviews is voluntary. You have the option at any time to cancel an interview and withdraw your consent to a recording and transcription of the interview without incurring any disadvantages.

**Data Protection Note**

By signing the consent form, you agree for members of the study team to collect and process your personal data, for the purposes of the aforementioned study. The principal investigator, Prof. Dr. Dr. Daniel Strech, is the responsible party for data processing according to the EU General Data Protection Regulation (GDPR).

The pseudonymized research data as well as the identifying data will be separately stored and processed on Charité drives, conforming to the GDPR and the Berlin Data Protection Law. Only members of the study team will have access to these data. After the retention period of 10 years (according to good scientific practice guidelines) has ended, we will delete or anonymize the identifying data. Please not that from then on, no withdrawal of consent, no information, change or deletion is possible, since we cannot connect you to the data.

You have the right to receive information (including a free copy) about all of your personal data from the principal investigator. You also have the right to withdraw your consent to the data processing at any time; in the case of withdrawal, you can also ask for deletion of your personal data. Please note that the legality of the data processing that has happened until then is left untouched: i.e., those data that have already been used for scientific publications are not subject to the withdrawal. To exercise these rights, please contact the principal investigator under the contact details below.

**Responsible Principal Investigator:** Prof. Dr. Dr. Daniel Strech, QUEST Center for Transforming Biomedical Research, Berlin Institute of Health, Translational Research Unit of the Charité – Universitätsmedizin Berlin, Charitéplatz 1, 10117 Berlin, Germany, phone: +49 30 450 543 068, email: , website: [www.charite.de](http://www.charite.de)

**Data Protection Officer:** You can always direct questions about the storage and processing of your data, or about your rights for data protection, to the Data Protection Officer of the Charité: Charité – Universitätsmedizin Berlin, Data Protection Office, Frau Janet Fahron, Charitéplatz 1, 10117 Berlin, phone: +49 30 450 580 016,

**Berlin Representative for Data Protection and Freedom of Information**: You have the right to complainat the Berlin Representative for Data Protection and Freedom of Information, which is the supervisory body for data protection. You have the right to veto at the responsible authority if you are under the impression that your data are being processed illegally: Berlin Representative for Data Protection and Freedom of Information (Berliner Beauftragte für Datenschutz und Informationsfreiheit), Friedrichstr. 219, 10969 Berlin, phone: +49 30 13889-0,

**Declaration of consent**

I hereby consent to participate in a semi-structured interview as part of the BRAVO project. I understand that my personal data will be collected, recorded, stored and processed for the purpose of the aforementioned study in a pseudonymized way. I was explained the type and aims of the study. I understand that the results of the study are published in an anonymized form, which does not allow for my direct identification. I had ample opportunity to ask the study team questions. I am aware that my participation is voluntary and that I, at any time, have the right to withdraw my consent without providing any reasons and without any disadvantages for myself, and that I can withdraw my consent to further processing of my data, as well as ask for their destruction.

__________	____________	___________________________________________

NAME	DATE	SIGNATURE

Link to dashboard + tutorial

Dear ________,

Thank you for your willingness to participate in the BRAVO interview study. Would any of the following days/times suit your calendar:

DATES/TIMES 
 
The interview will take place via Microsoft Teams. We will send you a reminder 1 day before the interview.

Please find the dashboard here (if the hyperlink does not work, copy-paste the following URL into your browser: [LINK]). You might want to watch this brief [tutorial](https://youtu.be/VDdljq5zI9E) where we explain how to navigate the dashboard. The dashboard is still under development, please do not share this link with others.

We look forward to talking with you on.

Kind regards,

Tamarinde Haven

**Appendix 2.** Proof-of-principle dashboard (in screenshots)

Screenshot 1. Landing page with explanation

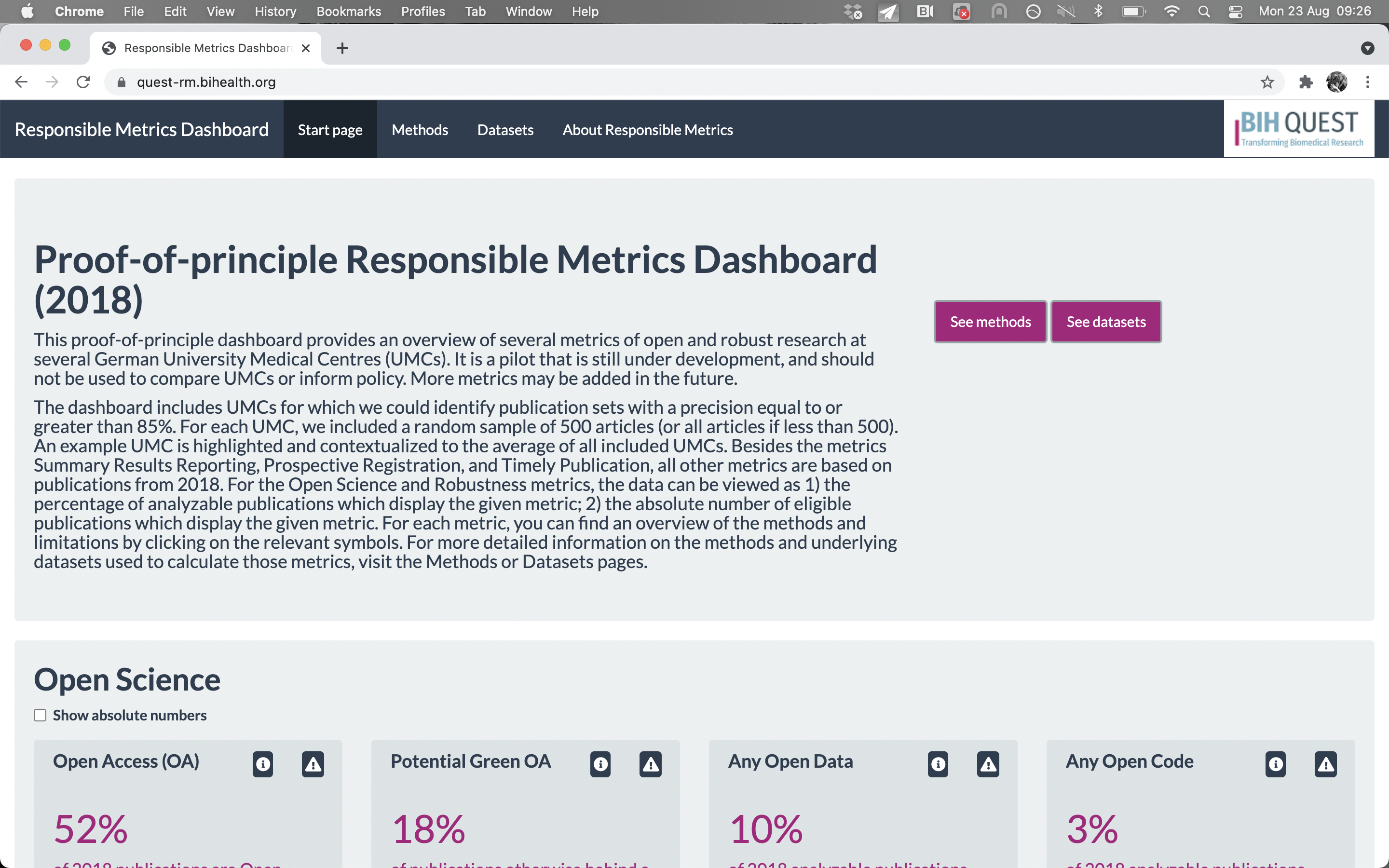

Screenshot 2. Metrics related to Open Science

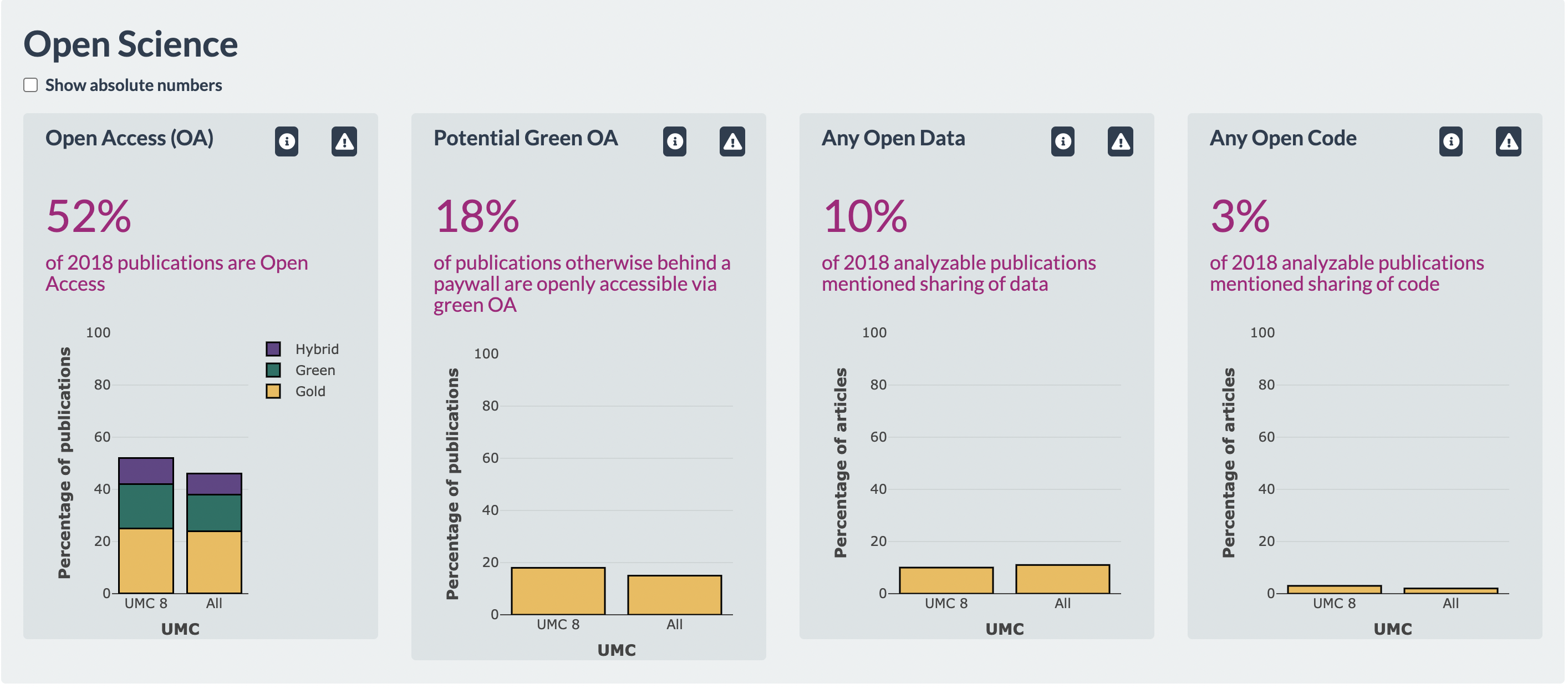

Screenshot 3. Metrics related to clinical trials

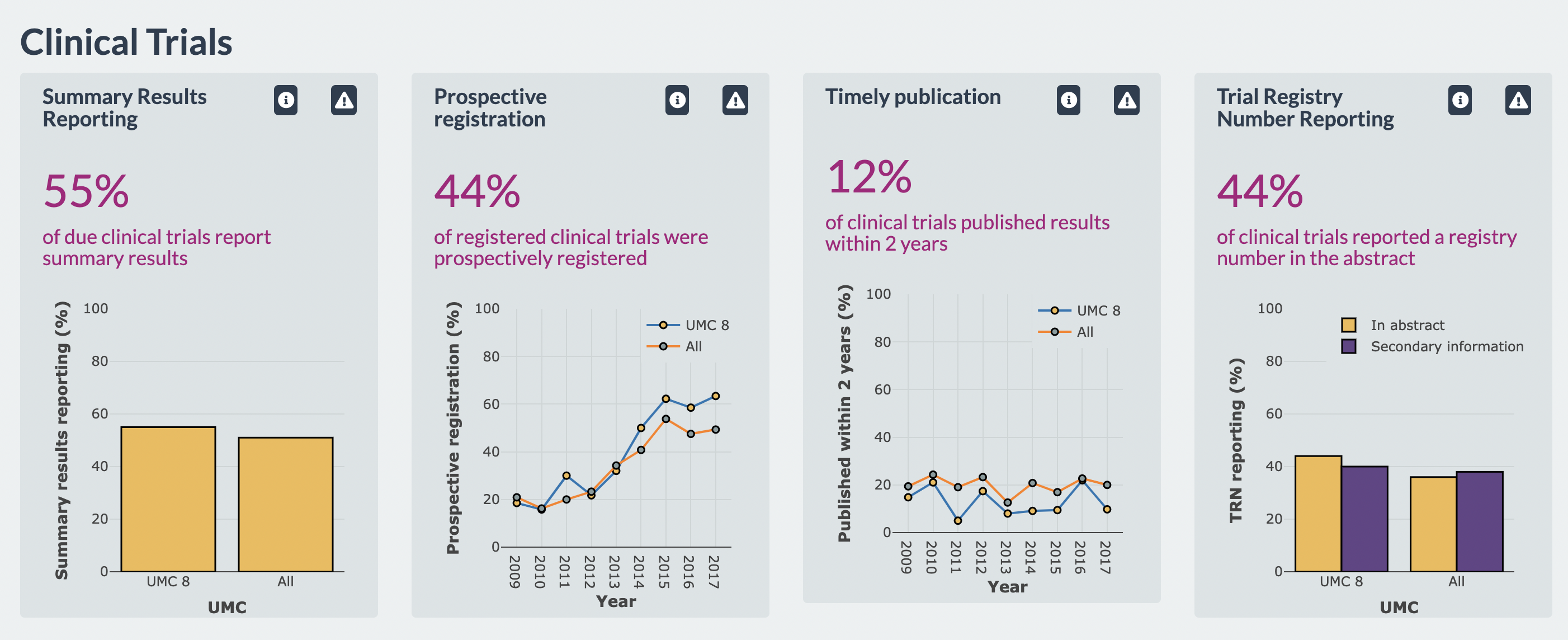

Screenshot 4. Metrics related to animal research

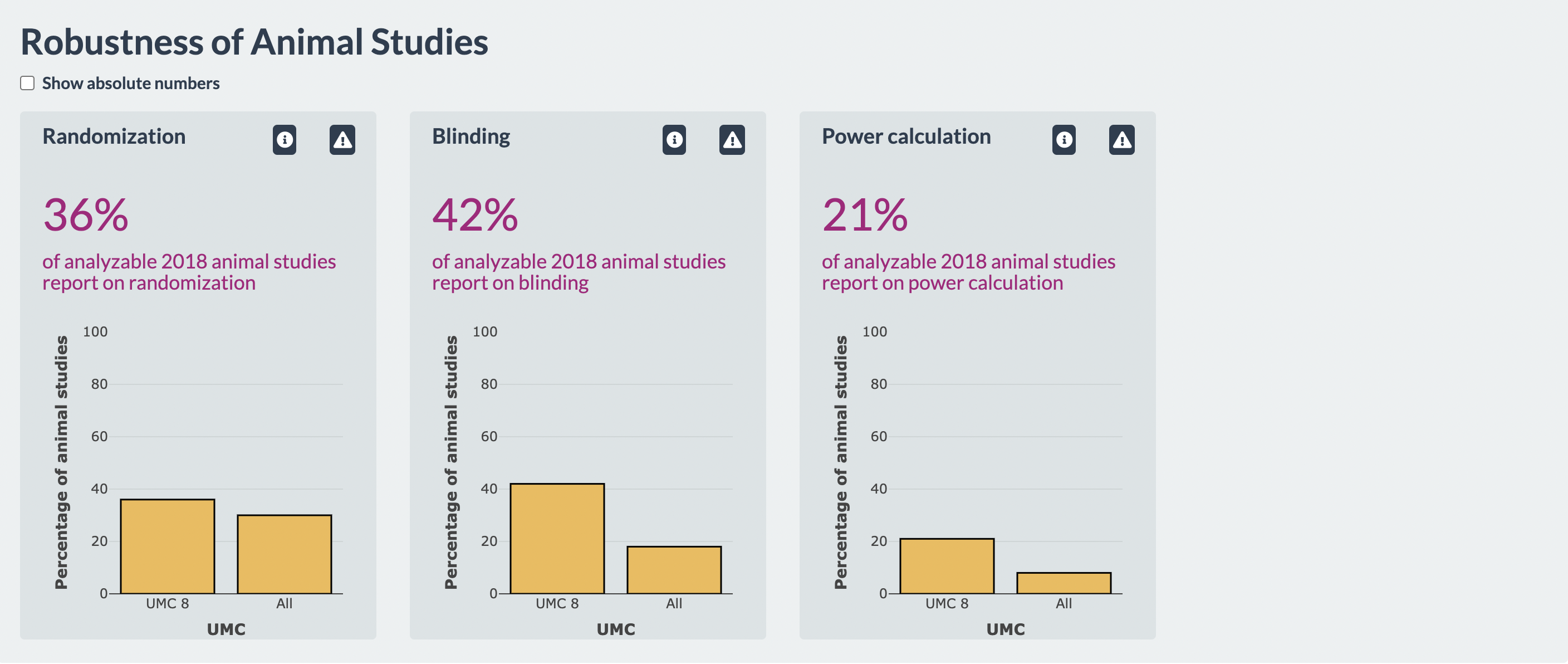

Screenshot 5. Demonstration “info icon” and “warning icon

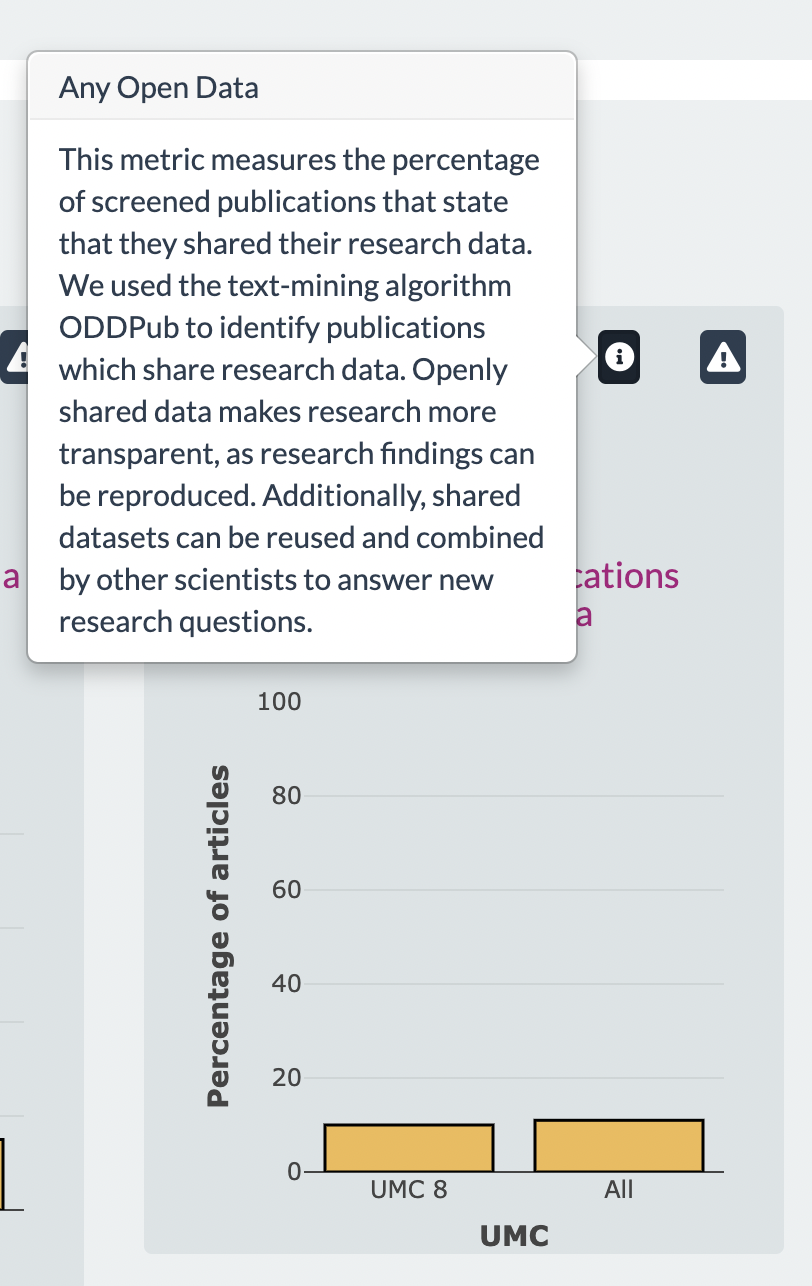

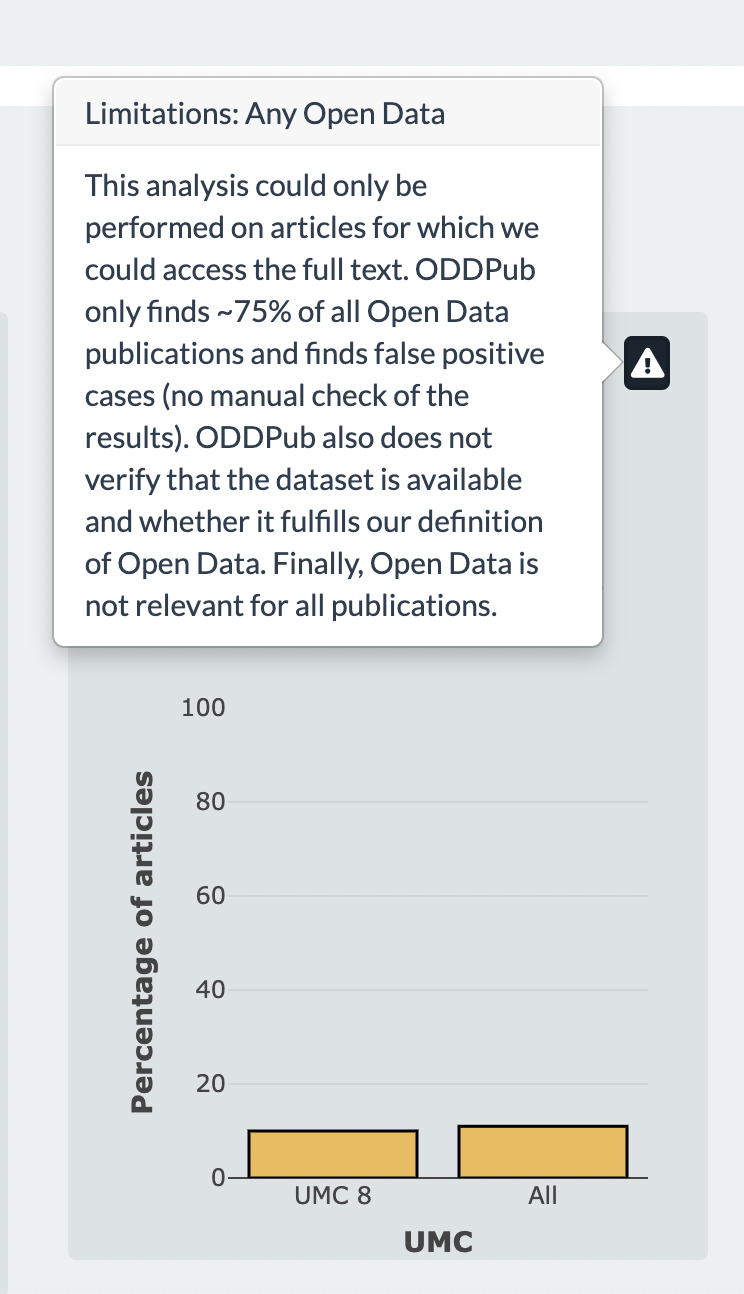

**Appendix 3.** Topic guide
Please note: *Italics* are alternative phrasings.

### Topic guide -- SWOTs of Dashboard approach.

#### Introduction interview

1. Open interview [“Thank you for participating in the interview”]

2.  Notify about recording [“I would like to emphasize that we record this interview, we will then transcribe the interview anonymously and after that the recording will be safely deleted.”]

3. Explanation of what will happen with the results [“The results will be used in a scientific paper, where we will illustrate our findings with quotes from the interview. These quotes are anonymized, but you might be able to recognise your own citations. However, nobody else should be able to.”]

4. Check informed consent [“Thank you for signing the informed consent form. I would like to repeat that participation is voluntary, that you can withdraw at any time without suffering any disadvantage. Do you have any questions before I start the recording?”]

5. Introduce yourself and themselves (name, position, institute) [“My name is TH and I work as postdoctoral researcher for QUEST that strives to increase the value of biomedical research at BIH and beyond. Could you perhaps introduce yourself?”]

6. Introduction project/goal interview: “We invited you to take part in these interviews that are part of the BRAVO project. The overarching goal of the BRAVO project is to increase the application of practices for robust and useful research across German University Medical Centers (UMCs). To realize this overarching goal, we are developing a dashboard that would allow UMCs to visualize the adoption of these responsible research practices by means of ‘metrics’ that relate to open science, timely reporting of clinical trials and robustness in animal research. The dashboard ‘dummy’ that you have been sent and are seeing now is our proof-of-principle version thereof. The goal of the dashboard is to provide institutions and other interested parties with a baseline indicator of the degree to which several practices for robust and open science are being performed at one specific institution. We invited you because we are very interested in your views on this dashboard and the metrics it includes. The goal of the interviews is for us to learn about the strengths and weaknesses of this dashboard and the metrics it includes, and to identify potential improvements. Do you have any other questions before we start?”

#### Core interview: SWOTs of dashboard

**‘Grandtour’ question:**
What is your view on a dashboard using these novel metrics?

**Deepening questions:**

What would you consider the strengths and weaknesses of such a dashboard?

*What do you consider possible pitfalls of such a dashboard?*

*What is the added value of such a dashboard?*

*Based on the dashboard, do you feel you have a good understanding of the*	*limitations of metrics displayed?*

How would you, in your institution, use such a dashboard?

*What would be, in your opinion, incorrect usage of such a dashboard?*

*What would/could you use such a dashboard for?*

*How should university leadership use such a dashboard?*

*What would be the possible (positive/negative) consequences of using this dashboard?*

*How could the information in this dashboard be abused, or what would be ‘gaming’ the information in this dashboard?*

*One foreseeable usage of this dashboard is that it could be used for benchmarking institutions, what are your thoughts regarding benchmarking?*

Which metric would you find most informative to support decision making at your institution?

* *How could this metric play a role?*

Which metrics do you believe should receive most attention and why?

#### Refinement and uptake questions

How should this dashboard be refined to promote uptake among UMC leadership?

*What suggestions do you have to further refine this dashboard?*

*Which feature of the dashboard should be improved (and how/why?)?*

*How could we optimise the dashboard to increase the chances of uptake among UMC leadership?*

**Snowballing** (optional)

With which other experts in your network should we talk to in order to better understand the situation?

#### Ending the interview

Is there anything else you’d like to say?

Can I contact you in case I need any additional information or if something is unclear?

Can we send you a summary of this interview with the option to comment or send corrections? (Member-checking)

Close the interview and thank the interviewee for their participation.

Code Tree

*Note:* This code system consists of three levels. The SWOTs in bold (e.g., “Strengths”, highest level), the themes in *italics* (e.g., “data and behavior driven” medium level – different topics that could be interpreted as SWOTs), and subcodes in regular font (e.g., “pick a core set of metrics”, lowest level – subcomponents of a theme).

**Code System**

**Strengths**

*Seeing where you stand*

Data and behavior driven

Creating a baseline

self-evaluation

*novel and relevant*

innovative and discipline-specific

alternative to current indicators

novel and timely

*clear presentation*

percentages and absolute numbers

interactive features

**Weaknesses**

*lack of clear overall framework*

not properly embedded in the policy landscape

infrastructure needs to be in place first

metrics do not represent robustness and transparency

lack of reasoning behind the metrics

*possibly outdated / no comparison over time*

attention for Open Science increased

*sample of publications*

better to use population

risk of sample-specific biases

*methods and conceptualization difficult to understand*

additional layer of data

unclear denominator

not all data sources covered

data sources and missingness not covered

metrics badly explained and operationalized

lack of consensus about terminology

qualifiers unclear (e.g., “Any…”)

**Opportunities**

*conversation starter*

fuel discussion how to improve

more holistic evaluation

add information how to induce change

induce collaboration or healthy competition

*benchmarking over time*

benchmarking against yourself

tracking progress

*internal usage only*

roll-out only after some delay

institutions choose what to make public

only the UMC leadership

*tailoring dashboard*

allow institutions to comment/correct the numbers

more fine-grained indicators

pick and choose indicators relevant indicators

center level or research field level

*complementing the dashboard with other indicators*

Open Science

diversity

preprints

General indicators

diversity

societal value and uptake of research

animal research

choice of model system

preregistration of animal research

clinical research

number of trials

patient engagement

turn dashboard into repository

*communication of metrics performance*

sensitivity and specificity

enrich information conveyed, e.g. with error bars

information on dashboard updates

**Threats**

*putting institutions in a bad light*

correct the records

not to be blamed for lack of infrastructure

*incorrect interpretation*

dynamic information

choice of comparator

numbers taken at face value

lack of context

*gaming metrics*

numeric indicators always wrong

manipulating information

Goodhart’s law

1. Macleod, M. R., Michie, S., Roberts, I., Dirnagl, U., Chalmers, I., Ioannidis, J. P. A., Salman, R. A.-S., Chan, A.-W., & Glasziou, P. (2014). Biomedical research: Increasing value, reducing waste. *The Lancet*, *383*(9912), 101–104.
2. Ioannidis JPA (2014) How to Make More Published Research True. PLOS Medicine 11(10): e1001747.

   Moher, D., Bouter, L., Kleinert, S., Glasziou, P., Sham, M. H., Barbour, V., Coriat, A.-M., Foeger, N., & Dirnagl, U. (2020). The Hong Kong Principles for assessing researchers: Fostering research integrity. *PLOS Biology*, *18*(7), e3000737.
3. Begley, C. G., Buchan, A. M., & Dirnagl, U. (2015). Robust research: Institutions must do their part for reproducibility. *Nature*, *525*(7567), 25–27.

   Flier, J. (2017). Faculty promotion must assess reproducibility. *Nature*, *549*(7671), 133–133.

   Strech, D., Weissgerber, T., & Dirnagl, U. (2020). Improving the trustworthiness, usefulness, and ethics of biomedical research through an innovative and comprehensive institutional initiative. *PLOS Biology*, *18*(2), e3000576. https://doi.org/10.1371/journal.pbio.3000576
